# Process evaluation of a New psychosocial goal-setting and manualised support intervention for Independence in Dementia (NIDUS-Family)

**DOI:** 10.1101/2024.02.06.24302127

**Authors:** Danielle Wyman, LT Butler, Sarah Morgan-Trimmer, Peter Bright, Julie Barber, Jessica Budgett, Kate Walters, I Lang, P Rapaport, Sara Banks, Marina Palomo, Vasiliki Orgeta, Gill Livingston, K Rockwood, K Lord, J Manthorpe, B Dow, J Hoe, Claudia Cooper

## Abstract

**Introduction:** We report a process evaluation embedded within a UK Randomised Controlled Trial (RCT), which demonstrated that New Interventions for independence in Dementia Study (NIDUS)-Family (a manualised, multimodal psychosocial intervention), was effective relative to usual care, on the primary outcome of Goal Attainment Scaling (GAS) over one year. We aimed to test and refine a hypothesised theory of change model delineating key causal assumptions for impact mechanisms.

**Methods:** In 2021-22, intervention-arm dyads completed an acceptability questionnaire developed to test causal assumptions. We interviewed dyads and their intervention facilitators, purposively selected for diverse follow-up GAS scores and sociodemographic diversity. Matching observational data were collected from intervention session recordings, using a checklist developed to test causal assumptions. We thematically analysed data, then integrated qualitative and quantitative data.

**Results:** 174/204 (85.3%) dyads allocated to NIDUS-Family, fully completed the intervention, 18 partially completed it, while 12 received none. 47/192 (24.5%) of carers receiving any sessions completed the acceptability questionnaire. 27/58 (47%) dyads purposively selected, and 9/10 facilitators participated in qualitative interviews; and we observed 12 sessions. We identified four themes: A) ‘Someone to talk to helps dyads feel supported’; B) ‘NIDUS-Family helps carers change their perspective’; C) ‘Personalisation helps people living with dementia maintain their identity’; and D) ‘Small steps help dyads move forward’.

**Conclusion:** Key causal pathway mechanisms were: regular sessions with a consistent facilitator providing space to discuss priorities, supporting carers to consider new perspectives and approaches to personalising care; and planning small actionable steps towards goals. Findings will support NIDUS-Family implementation.

## INTRODUCTION

Around 944,000 people live with dementia in United Kingdom (UK). Only two-thirds have a diagnosis (1), of whom under half receive post-diagnostic support (2). Over 700,000 unpaid family carers (henceforth carers) provide support, many reporting high rates of distress and morbidity (3). National Health Service England’s Well Pathway for Dementia and other initiatives recommend routinely offering post-diagnostic care (4).

Structured psychosocial interventions can be facilitated by trained, supervised staff without clinical qualifications, increasing access to evidence-based care. A new manualised, modular post-diagnostic support intervention (NIDUS (New Interventions for Independence in Dementia Study) – Family), delivered remotely by non-clinical facilitators, improved attainment of carer-rated personalised goals for people living with dementia in their own homes, relative to goal-setting alone in a Randomised Controlled Trial (RCT) (5). NIDUS-Family is the first fully manualised intervention tailored to individualized goals dyads (person and carer) set, by selecting modules involving behavioural management, psychoeducation, communication skills, enablement and environmental adaptations (6,7).

Process evaluations provide an understanding of interventions to inform future implementation (8). We aimed to test and refine the NIDUS-Family theoretical model of change (Figure 1) to identify factors influencing how it was implemented in the trial, and mechanisms and contextual factors influencing its effectiveness (9).

**Figure 1.**
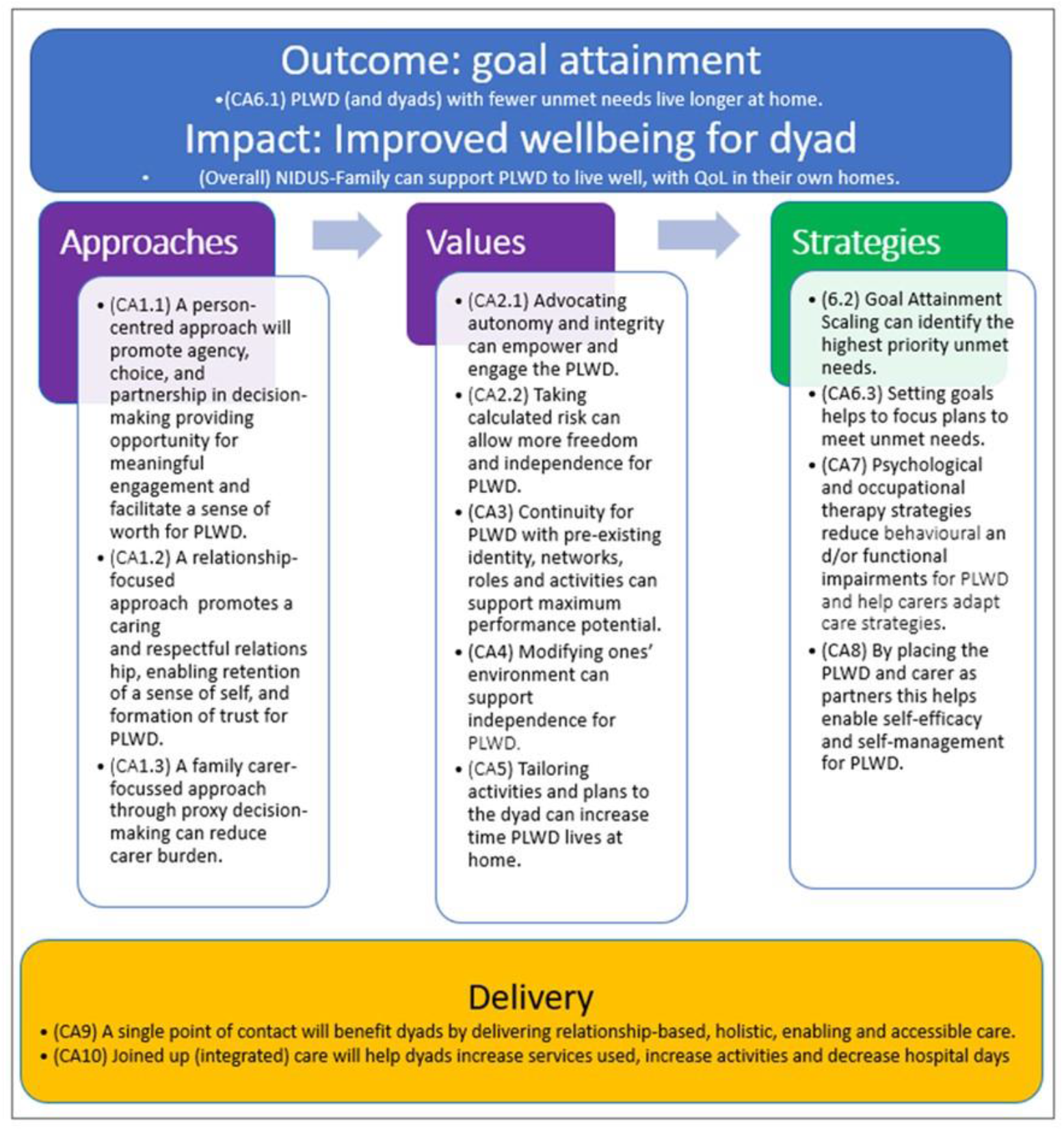
Initial theoretical model of change for attainment of dyadic goals through NIDUS-Family intervention (with associated causal assumptions) (9,10)

## METHODS

### Design

We used a convergent mixed-methods design (10). The NIDUS-Family theoretical model (Figure 1) was pre-specified (10) and outlined causal assumptions regarding how the intervention might effect change, which interview questions and analyses were designed to test. London-Camden and King’s Cross National Research Ethics Committee (19/LO/1667) approved the study; its protocol is registered (ISRCTN11425138) and published (10).

### Setting and sample

We obtained (written or audio-recorded) informed consent from all dyads allocated to the NIDUS-Family intervention arm. Dyads were purposively sampled for diversity in demographic characteristics (gender, relationship between dyad) and 12-month family-carer rated GAS (Goal Attainment Scale) scores. Selected dyads, and all intervention facilitators, were invited to be interviewed and their available recorded intervention sessions observed. Facilitators were asked to record at least one session per dyad. Sample size was planned to ensure sufficient diversity of cases (11).

### Intervention description

NIDUS-Family was delivered by university-employed facilitators, who were psychology or social science graduates without clinical training or qualifications. Facilitators were trained, and their competency assessed by role plays. They attended fortnightly group supervision with a clinical psychologist. NIDUS-Family comprised 6-8 manualised sessions over six months, by video-call/telephone (in-person when Covid-19 restrictions permitted). Sessions included dyads, or just the carer where this was agreed between facilitator and carer/ dyad to be most appropriate. These sessions were followed by 30-minute catch-up telephone/video calls at 2– 3-month intervals (dyads preference), between 6-12 months from baseline to review progress towards goals, following a standard guide. Full adherence was defined as receiving 6-8 sessions, including the final session, which consolidated learning across the intervention to develop an action plan. Further details are published (5).

### Data collection

Socio-demographic and attendance data were collected (see protocol (10)). After 12-month follow-up, all intervention-arm carers were invited to complete via post or telephone, at their preference, a mixed methods ‘acceptability’ questionnaire’ (10), which we designed to test causal assumptions (Figure 1).

With purposively selected dyads (carers only where the person with dementia was unable to take part), in 2021-2, a researcher (DLW) conducted:

- Qualitative semi structured video/phone interviews, with dyads (or carer only) and facilitators (see (10) for topic guides), exploring experiences of NIDUS-Family. Interviews were audio-recorded, anonymised, transcribed verbatim and organised using NVivo V.12.
- Observations, by listening to at least one available recorded interview session for interviewed dyads. A checklist captured qualitative (‘free-text’) and quantitative (Likert scale) data relating to emerging theory, as the protocol describes (10).

### Analysis

We described intervention adherence, ratings of acceptability, and observational checklist quantitative data using summary statistics; thematically analysed (using reflexive methods) qualitative dyadic and facilitator interviews, observational checklist and acceptability questionnaire free text data (12), then integrated quantitative and qualitative data using a joint display, mapping findings to consider whether they supported causal assumptions (10).

## RESULTS

### Quantitative findings

Table 1 displays socio-demographic characteristics of the 204 trial intervention arm participants. Participants received the intervention face-to-face (n=3), by phone (n=63) or via video (n=126) (5).

**Table 1.**
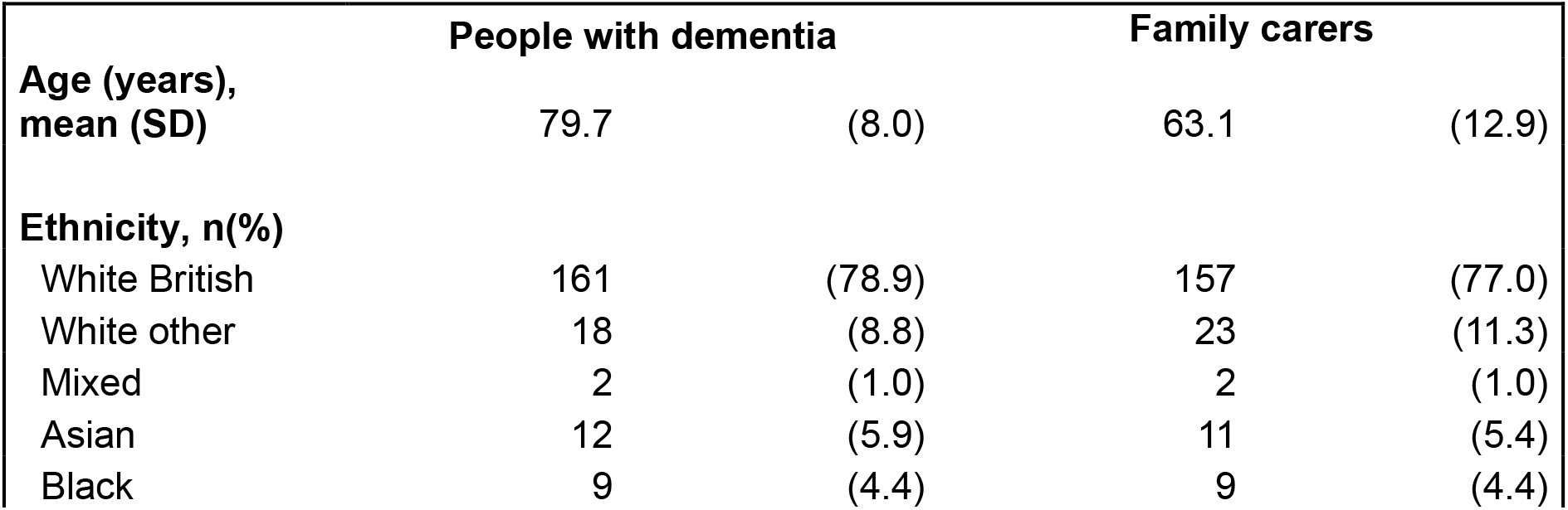
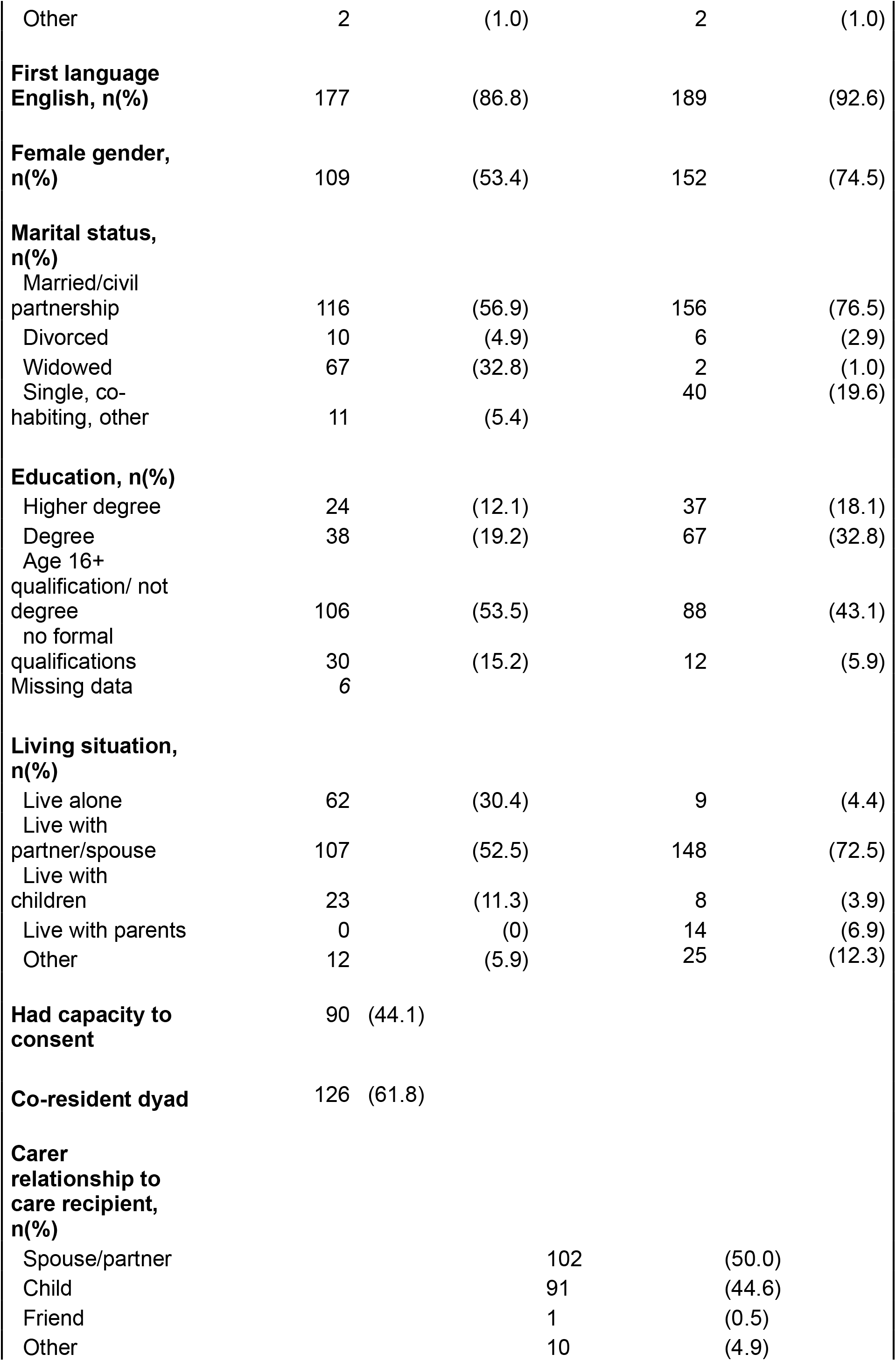
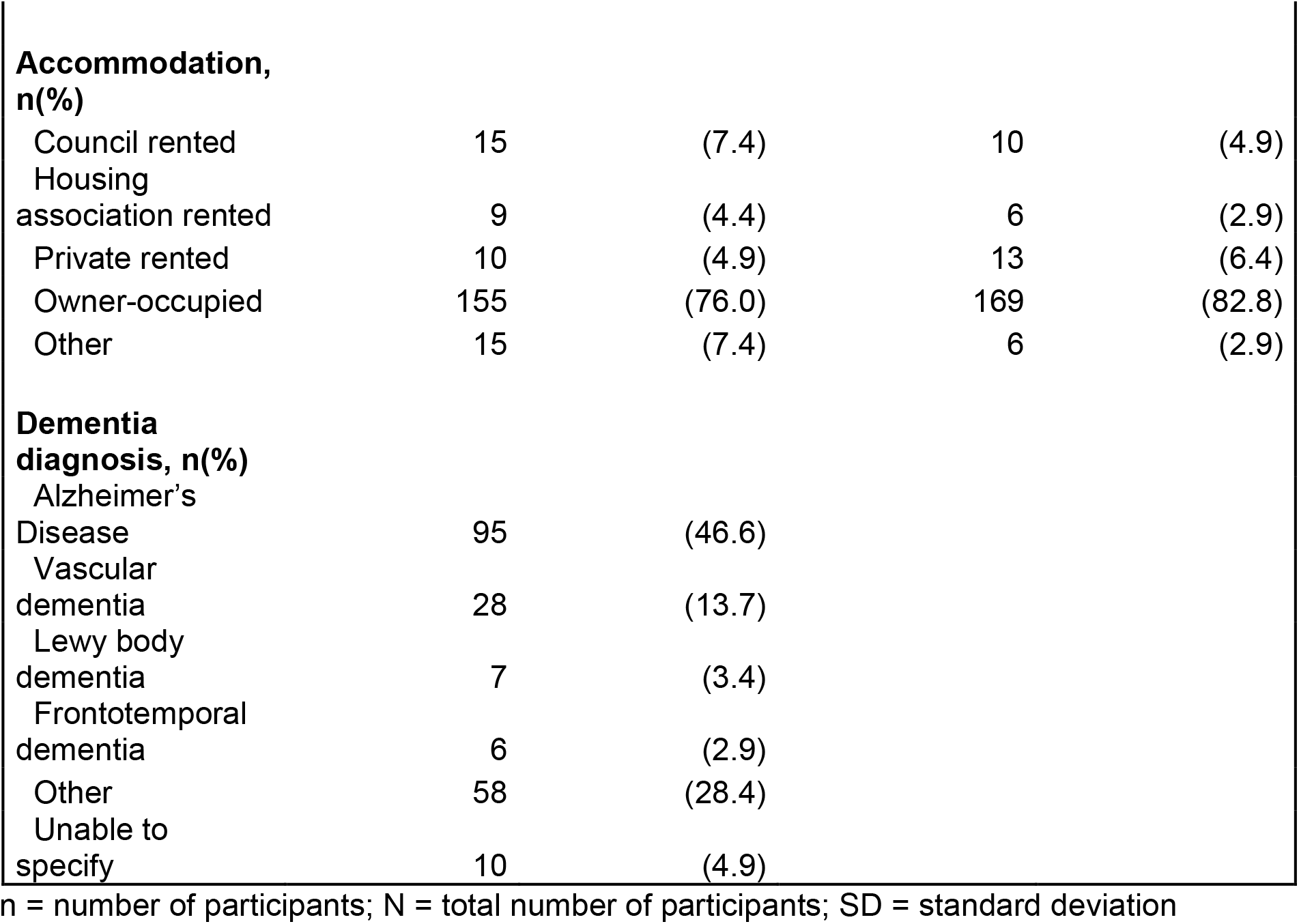
Baseline characteristics of dyads randomised to the NIDUS-Family intervention arm (n=204, base population for process evaluation)

#### Adherence

174 (85.3%) dyads fully (see methods for definition), and 18 partially completed the intervention; 12 received no sessions. Reasons for non-completion were: person living with dementia died (n=5), moved to a care home, (n=5), was hospitalised (n=1), became unwell (n=2); their carer died (n=1), or became unwell (n=2). Eleven dyads stopped because they found it not beneficial (n=3), lacked time (n=2), experienced family conflict (n=1), found discussions upsetting (n=1), or without giving a reason (n=3). Three dyads became uncontactable. Compared with the whole sample, fewer non-completing dyads were co-resident (50%, n=15), married/partners (36.7%, n=11), or included a care recipient with capacity to consent (30%, n=9).

#### Acceptability questionnaires

47/139 (34%) distributed acceptability questionnaires were returned [47/192, 24.5% of those receiving any intervention]. None were from non-completers. Table 2 shows characteristics of carers completing it (D27-65), of whom eight were also interviewed. In 21 (44.7%) dyads returning questionnaires, the care recipient had capacity to consent, and 26/44 (59.1%) were married/partners. Carers reported mainly positive experiences of the intervention (Figure 2).

**Table 2.** Matrix showing data available for 65 dyads who contributed data to the process evaluation.

| Dyad ID | Relationship (FC/ PLWD) | PLWD Capacity to consent to interview | Facilitator number | Goal set with | Dyad Interviewed | Session observation complete Yes/Y or No/N | AQ completed |
| --- | --- | --- | --- | --- | --- | --- | --- |
| D1 | Daughter/ Mother | N | F8 | FC | Y | Y | Y |
| D2 | Wife/ Husband | N | F1 | FC | Y | N | N |
| D3 | Wife/ Husband | Y | F9 | FC & PLWD | Y (FC&P LWD) | N | N |
| D4 | Son/Father | N | F6 | FC | Y | Y | Y |
| D5 | Daughter/<br>Mother | Y | F9 | FC | Y | Y | N |
| D6 | Wife/<br>husband | Y | F9 | FC &<br>PLWD | Y<br>(FC&P<br>LWD) | N | N |
| D7 | Daughter-<br>in-law/Mother<br>-in-law | Y | F1 | FC &<br>PLWD | Y | N | Y |
| D8 | Wife/<br>Husband | Y | F2 | FC | Y | Y | N |
| D9 | Daughter/<br>Father | Y | F9 | FC | Y | N | N |
| D10 | Daughter/<br>Mother | N | F9 | FC | Y | Y | N |
| D11 | Daughter/<br>Mother | Y | F2 | FC | Y | Y | N |
| D12 | Daughter/<br>Mother | N | F3 | FC | Y | N | N |
| D13 | Wife/<br>Husband | N | F8 | FC | Y | Y | Y |
| D14 | Wife/<br>Husband | N | F2 | FC | Y | Y | N |
| D15 | Daughter/<br>Mother | N | F5 | FC | Y | N | N |
| D16 | Husband/<br>wife | N | F8 | FC | Y | N | N |
| D17 | Husband/<br>wife | Y | F3 | FC | Y | N | N |
| D18 | Wife/<br>Husband | N | F2 | FC | Y | Y | Y |
| D19 | Daughter/<br>Father | Y | F8 | FC | Y | N | Y |
| D20 | Wife/<br>Husband | N | F10 | FC | Y | N | N |
| D21 | Husband/<br>Wife | Y | F10 | FC &<br>PLWD | Y<br>(FC&P<br>LWD) | Y | N |
| D22 | Daughter/<br>Mother | N | F8 | FC | Y | N | Y |
| D23 | Wife/<br>Husband | N | F2 | FC | Y | Y | Y |
| D24 | Wife/<br>husband | N | F6 | FC | Y | Y | N |
| D25 | Daughter/<br>Father | N | F10 | FC | Y | N | N |
| D26 | Wife/<br>husband | N | F6 | FC | Y | N | N |
| D27 | Wife/<br>Husband | Y | F10 | FC &<br>PLWD | Y<br>(FC&P<br>LWD) | N | Y |
| D28 | Son/Father | Y | F2 | FC | N | N | Y |
| D29 | Son/<br>Mother | N | F2 | FC | N | N | Y |
| D30 | Daughter/<br>Mother | N | F2 | FC | N | N | Y |
| D31 | Wife/<br>Husband | N | F6 | FC &<br>PLWD | N | N | Y |
| D32 | Daughter/<br>Mother | N | F6 | FC | N | N | Y |
| D33 | Wife/<br>Husband | N | F6 | FC | N | N | Y |
| D34 | Daughter/<br>Mother | Y | F8 | FC &<br>PLWD | N | N | Y |
| D35 | Daughter/<br>Mother | N | F6 | FC | N | N | Y |
| D36 | Daughter/<br>Mother | N | F6 | FC | N | N | Y |
| D37 | Partner | Y | F6 | FC | N | N | Y |
| D38 | Partner | Y | F6 | FC | N | N | Y |
| D39 | Wife/<br>Husband | Y | F6 | FC | N | N | Y |
| D40 | Daughter/<br>Mother | N | F6 | FC | N | N | Y |
| D41 | Wife/<br>Husband | Y | F4 | FC | N | N | Y |
| D42 | Husband/<br>Wife | N | F4 | FC | N | N | Y |
| D43 | Daughter/<br>Mother | Y | F2 | FC | N | N | Y |
| D44 | Wife/<br>Husband | Y | F4 | FC | N | N | Y |
| D45 | Daughter/<br>Mother | N | F4 | FC | N | N | Y |
| D46 | Wife/<br>Husband | Y | F8 | FC &<br>PLWD | N | N | Y |
| D47 | Friend/<br>Friend | Y | F6 | FC | N | N | Y |
| D48 | Wife/<br>Husband | Y | F6 | FC | N | N | Y |
| D49 | Wife/<br>Husband | Y | F6 | FC &<br>PLWD | N | N | Y |
| D50 | Partner | Y | F2 | FC | N | N | Y |
| D51 | Husband/<br>wife | N | F2 | FC | N | N | Y |
| D52 | Wife/<br>Husband | N | F2 | FC | N | N | Y |
| D53 | Wife/<br>Husband | N | F10 | FC | N | N | Y |
| D54 | Wife/<br>Husband | Y | F10 | FC &<br>PLWD | N | N | Y |
| D55 | Husband/<br>wife | N | F4 | FC | N | N | Y |
| D56 | Wife/<br>Husband | Y | F8 | FC &<br>PLWD | N | N | Y |
| D57 | Wife/<br>Husband | Y | F10 | FC | N | N | Y |
| D58 | Son/<br>Mother | Y | F4 | FC &<br>PLWD | N | N | Y |
| D59 | Son/Father | N | F4 | FC | N | N | Y |
| D60 | Wife/<br>Husband | N | F10 | FC | N | N | Y |
| D61 | Husband/<br>Wife | N | F10 | FC | N | N | Y |
| D62 | Wife/<br>Husband | Y | F10 | FC | N | N | Y |
| D63 | Unknown<br>as Anon | ? | ? | ? | N | N | Y |
| D64 | Unknown<br>as Anon | ? | ? | ? | N | N | Y |
| D65 | Unknown<br>as Anon | ? | ? | ? | N | N | Y |

AQ: Acceptability Questionnaire; FC: Family carer; N/A not available; PLWD: Person Living With Dementia

**Figure 2.**
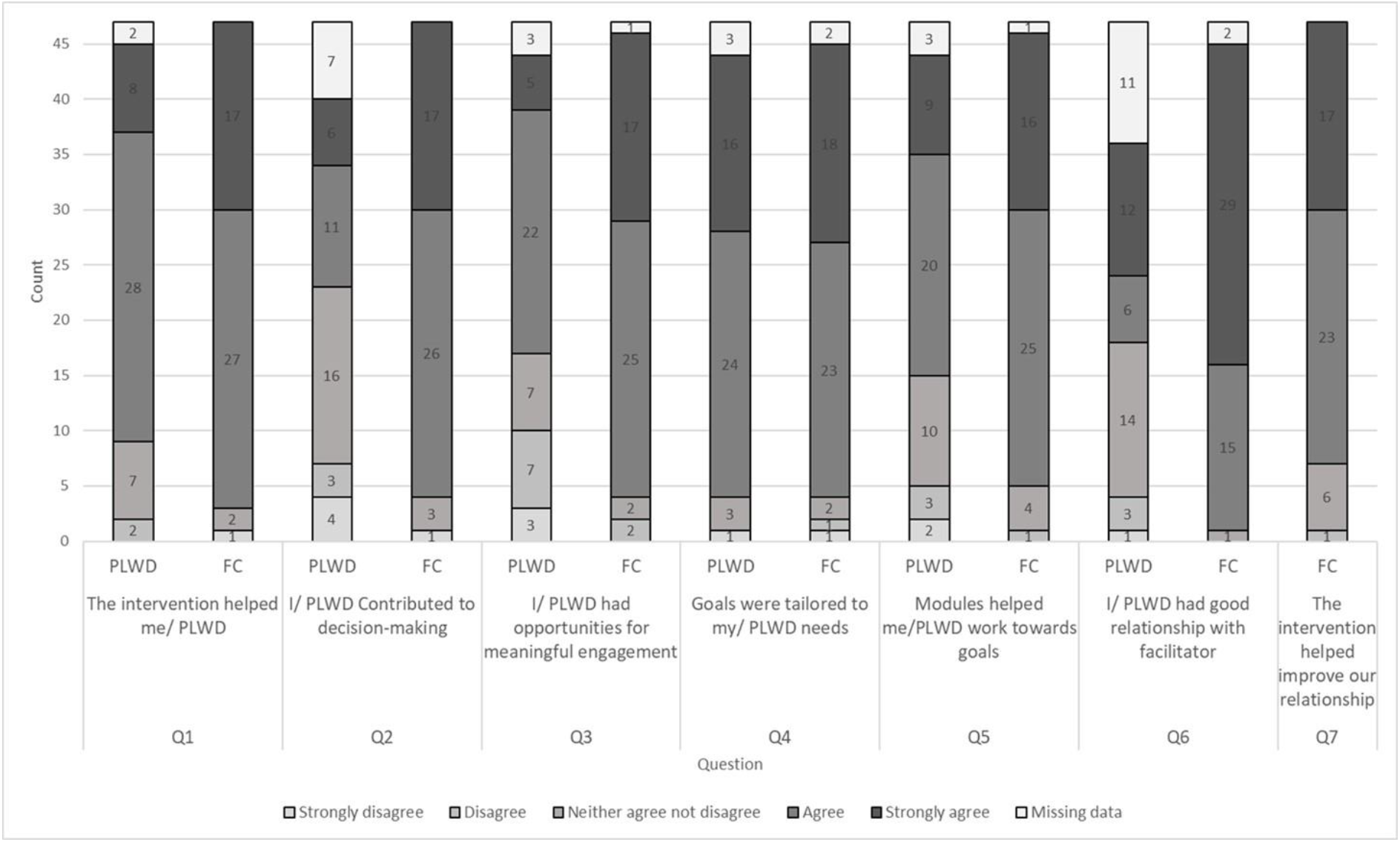
Acceptability questionnaire responses to discrete choice questions (n=47)

#### Session observations

As few sessions were recorded (due to client or facilitator preference), we only rated 12 recordings (one for 44.4% of interviewed dyads). Ratings indicated high fidelity to intended NIDUS-Family values and approaches, with statements relating to observing discussion around risks and setting clear objectives least often endorsed (Appendix, Figure 1S).

### Qualitative findings

27/58 (47%) of dyads approached took part in interviews (D1-D27; Table 2), as a dyad (n=4) or carer alone (n=23). Of these, 15 dyads were married (12 female and 3 male spouse carers), 19 were intergenerational (almost all mother/daughter). 9/10 facilitators (F1-F9) were interviewed.

We developed four themes: A) *‘Someone to talk to helps dyads feel supported*’ captured the value of a consistent space and facilitator, providing a third person perspective; B) *‘NIDUS-Family helps carers change their perspective’* described how NIDUS-Family supported carers to reflect and consider new perspectives and approaches; C) *‘Personalisation helps people living with dementia maintain their identity’* describes how sessions and goals were tailored to be meaningful and support personhood, though this was more challenging where the dementia led to severe impairment; and D) *‘Small steps help dyads move forward’* captured how the intervention supported steps towards goals.

### Theme A ‘Someone to talk to helps dyads feel supported’

Carers valued how NIDUS-Family provided space for discussions with a consistent facilitator, which enabled them to organise their thinking and make plans:

> *“I looked forward to it because I was able to talk to somebody*… *you just need to talk about all these things, because sometimes it’s a muddle in your head” (Interview, carer, D20)*

A positive relationship with the facilitator enabled dyads to openly “*talk about their issues*” (AQ, D13 (note all AQ quotes are carers)), “*difficulties and problems*” (AQ, D49). Dyads valued a facilitator who was “*collaborative*” (AQ, D30), “*supportive*” (AQ, D29), “*sensitive*” (AQ, D30), “*a good listener…followed up with [their] promises…was on time*” (AQ, D39), “*kind and understanding*” (AQ, D34). One person with dementia valued discussing concerns with “*someone … who actually knew what they were talking about… put some of our fears to the side” (Interview, person with dementia, D3)*.

Positioning the facilitator as an ‘*outsider’* encouraged discussion of issues carers felt unable to ‘*burden’* family with (Interview, carer, D23). A carer described how she was supported to take action:

> *“making the decisions that were worrying me, like the DNR [Do Not Resuscitate], I knew I had to do it but I was scared and she spoke to me … And then I said, could I make an appointment and I made it and I just wished I’d have done it a lot longer before.” (Interview, carer, D23)*

Observation notes described facilitator ‘*compassion’*, ‘*encouragement’* (D4, D10, D21) and ‘*support’* (D8, D24), ‘*empathetic’* and ‘*kind feedback’* (D1, D11, D14), support of the dyad’s ‘*challenges’* (D5, D14), ‘*positive reflection’* (D4, D8), and ‘*positive reinforcement’* (D8, D18, D21).

#### Theme B ‘NIDUS-Family helps carers change their perspective’

Sessions provided space to reflect, to “look objectively” (AQ, D31); together with increased understanding of the illness and symptoms, this enabled carers to reappraise their role and be *“less hard”* (AQ, D33) on themselves as carers. One carer explained how the intervention caused her to reflect on the importance of self-care:

> *“she [facilitator] made me start thinking about myself because my whole concern was about my husband. And she was the first one that made me realise that, actually, I’m very important myself and I’ve got to keep going myself to be able to look after him” (Interview, carer, D2)*

NIDUS-Family helped carers re-frame their approach to caring. Some carers described how previously they felt burdened by the need to *“carry on with [caring]”* (Interview, carer, D12), feeling left alone to *“just get on with it”* (Interview, carer, D23), and *“having no support*” (Interview, carer, D4) from family, friends, or local services. NIDUS-Family shifted this perspective, enabling carers to prioritise and organise their plans to meet care needs:

> *“Rather than just feel that you’re sort of lost, not doing enough or not doing anything or isolated, yes. So it [NIDUS] gave, yes, it gave us that ability to organise ourselves to deliver the care that Dad needs but also get on with our own lives.” (Interview, carer, D19)*

There was a sense of enablement in carers’ accounts. Facilitators offered carers alternative perspectives, and support to “*make difficult decisions*” (AQ, D23); *“take control of things”* (AQ, D1), and ask for help, as described in this next quote:

> *“And so I’ve got to learn to change that [asking for help], but it has made me braver. It has made me realise I have limitations, so it’s been useful.” (Interview, carer, D24)*

A carer described how developing a better understanding of the illness helped the family reconsider the care approaches likely to work best:

> *“…the main thing was the understanding of how my mum’s mood affected her and how she was and her behaviour. So for us to … understand that a bit more, we could deal with the whole situation in a different way.” (Interview, carer, D22)*

Dyads valued the quotes from carers in modules, which gave them confidence that the suggestions might help them, as they had others in a similar position:

> *“relief that I wasn’t alone …. I remember thinking, this isn’t going to work but I will give it a try because it worked for other people.” (Interview, carer, D1)*.

Observations captured how carers were “*proactive*” (D8, D14, 24) and “*took ownership*” (D14, D18, D21) by “*planning*” strategies or activities (D4, D8, D10, D21). Some carers and facilitators suggested it would be beneficial to set some goals during as well as before the intervention, so new perspectives that the intervention enabled could inform them:

> *“So maybe setting of goals … further down the line so that we can actually see … why we’re setting goals, and be more realistic about the goals we set.” (Interview, carer, D24)*

Supporting this, a facilitator described how when a goal appeared unrealistic, they used the sessions to support the carer to take a different perspective on what change was possible:

> *“the [carer] wanted her to communicate verbally …that she wanted to use the bathroom, for example, and I did set the goals at the time. But then when I had the first session I got to know a lot more … he had some unrealistic expectations and that what we could do with the intervention was to make him aware that communication, for example, is not only verbal but it’s also non-verbal.” (Interview, Facilitator 8)*

### Theme C ‘Personalisation helps people living with dementia maintain their identity’

This theme captures how dyads and facilitators tailored goals and sessions to be meaningful and feasible, and the challenges of doing so where people living with dementia were less able to be actively involved. Facilitators discussed how intervention flexibility enabled this:

> *“straight away from setting those goals around areas that were meaningful to them*

*…. so I think that it felt more personal to the dyad, it felt more individualized, and so it felt like you’re making kind of a real difference, rather than just kind of reading through material.” (Interview, Facilitator 2)*

Where the person with dementia was able to select their own goals, the process of supporting participants to choose activities of interest was more straightforward. In this next quote, a participant with dementia described choosing a goal to reflect a past interest. This process of rediscovering past interests supported the intended mechanism that NIDUS-family would help support independence by enabling people living with dementia to maintain or rediscover their identities:

> *“I chose it because I’ve always done woodwork … with getting back into it, well it’s*

*something I look forward to doing now, yeah.” (Interview, person living with dementia, D6)*

Where the person did not wish to plan goals and activities, there was a tension between encouraging this and respecting choice, as described in this next quote:

> *“I am trying to retain his independence and his choices as much as possible because it would be very easy to say, “Do this, do that, do the other,” but you can’t do that, you shouldn’t do that, let’s put it that way.” (Interview, carer, D24)*

A carer discussed how the initial goal set for the person they cared for was in hindsight, from her new perspective, informed by NIDUS-family discussions *“ridiculous”* (Interview, carer, D13) as her relative no longer could achieve it. One carer decided to do more of what their relative currently enjoyed, as they struggled to identify a new activity the person would be interested in and have capacity to start:

> *“So going to the hairdressers and having the foot lady, instead of doing those things once a month we now do them twice a month and we do it that way.” (Interview, carer, D5)*

#### Theme D ‘Small steps help dyads move forward’

Observations captured how NIDUS-Family involved co-creating manageable steps towards goals. Several carers also described how plans were informed by developing an in-depth understanding of the problems being addressed:

> *“if it had been a textbook I was reading or, you know, it wouldn’t have worked in quite the same way as just breaking it down to the minutia of relationship and what was going on in my life between my mum and for my mum” (Interview, carer, D5)*

Carers described how they were able to use this new understanding and work with facilitators to make plans, and break these down into small steps that made a difference:

> *“The intervention enabled us to really think about what the challenges and problems were which were impacting quality of life. Once focussed on this our researcher came up with excellent ideas that we could then act upon. These ideas and*

> *suggestions have improved my mother’s life immeasurably and without the intervention we would be stuck where we were -and even worse given the situation with COVID.” (AQ, carer, D65)*

This process of trying out new plans was iterative – learning from what did and did not work. The intervention structure enabled dyads to reflect on their goals and progress:

> *“Because we re-visit the goals frequently - she [mum?] can now remember what they are without any/few clues. So she keeps working on some of them.” (AQ, D64)*

### Data integration (Table 3)

The integrated analysis evidenced all causal pathways except CA4 and CA10 were implemented. It identified core mechanisms for goal attainment as: a consistent facilitator (CA9) delivering relevant approaches (CA1.1 and CA1.2 for people with dementia, CA1.3 for carers) and strategies (CA6.2, CA7, CA5) across regular sessions.

**Table 3.**
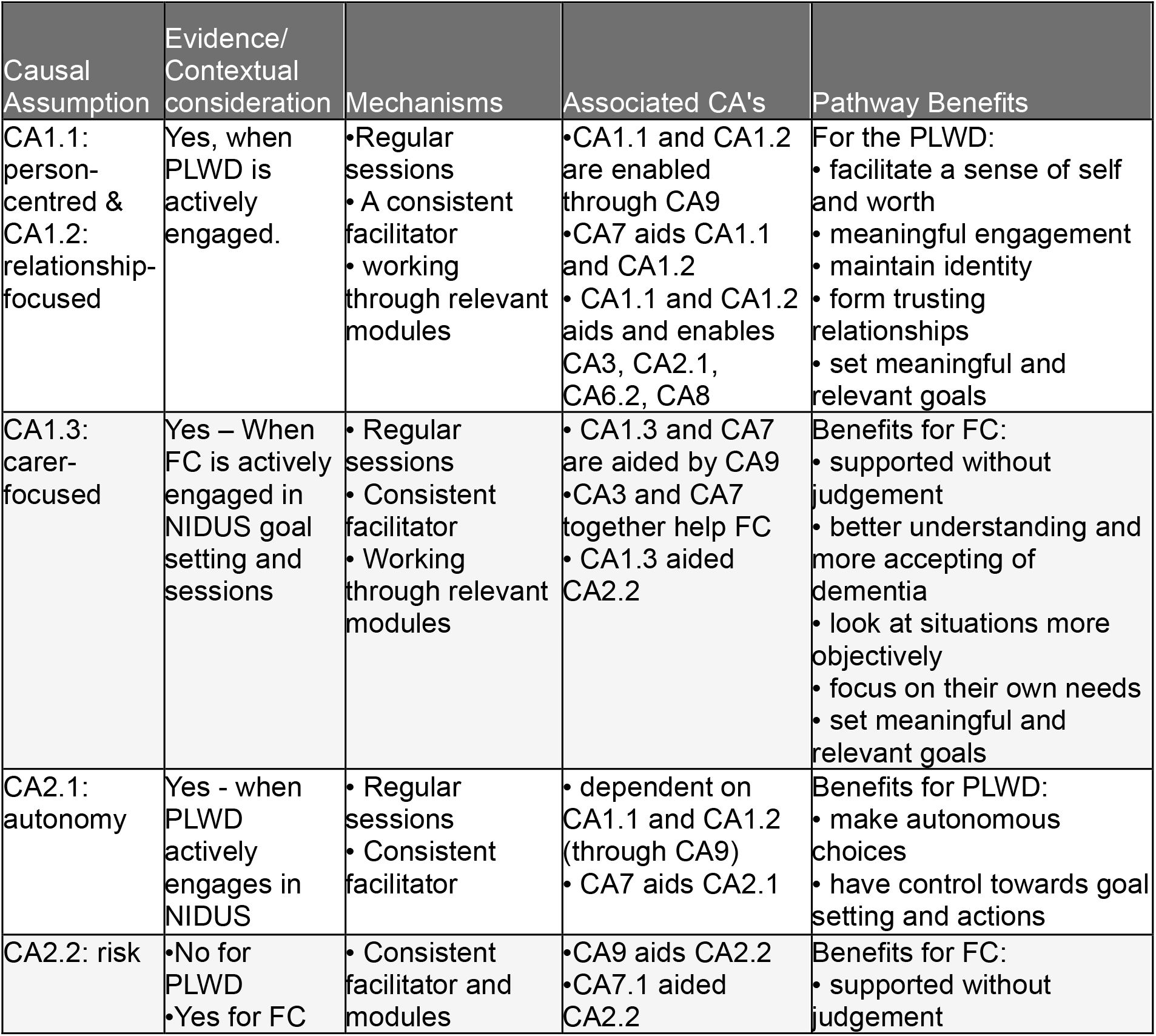
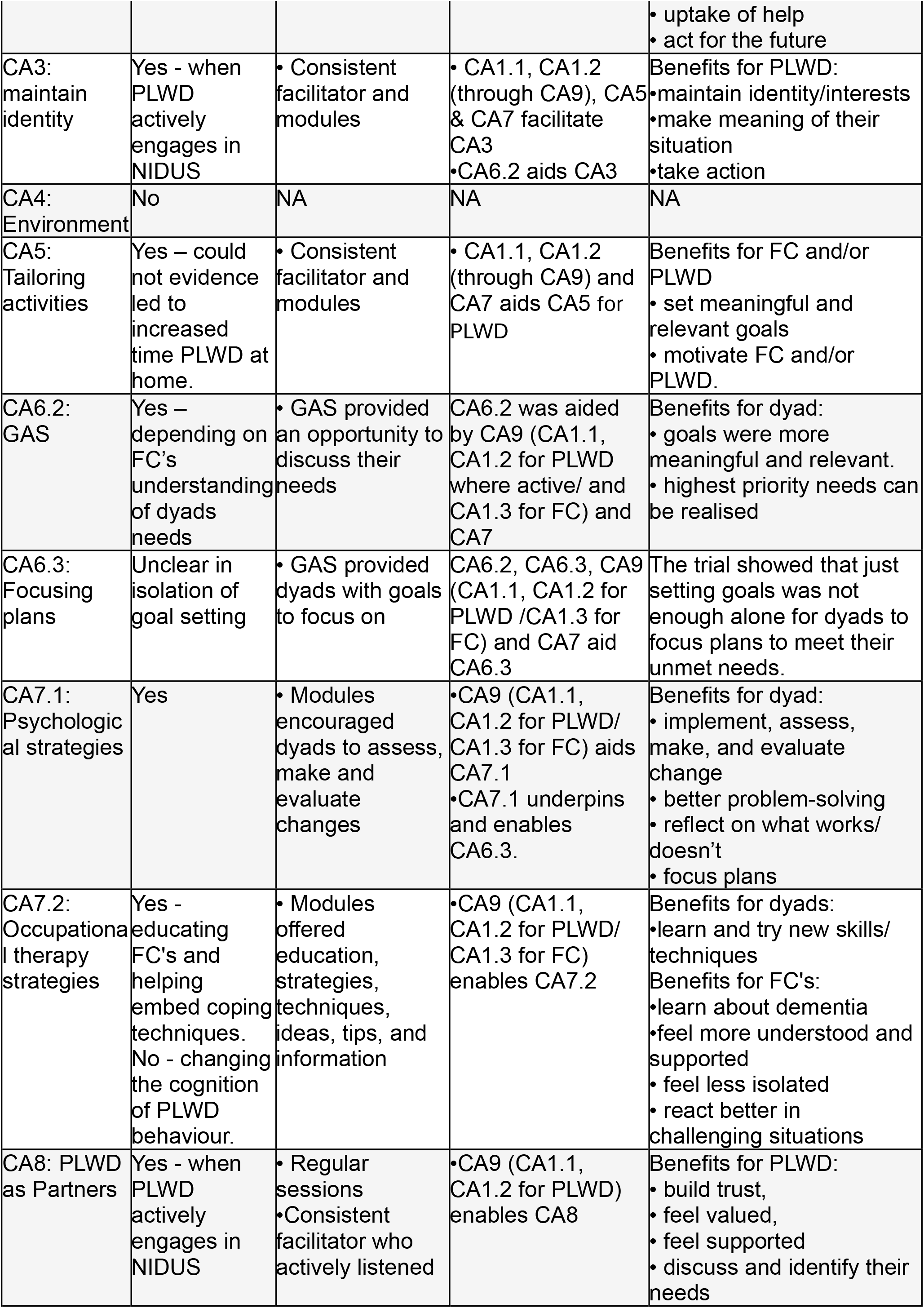
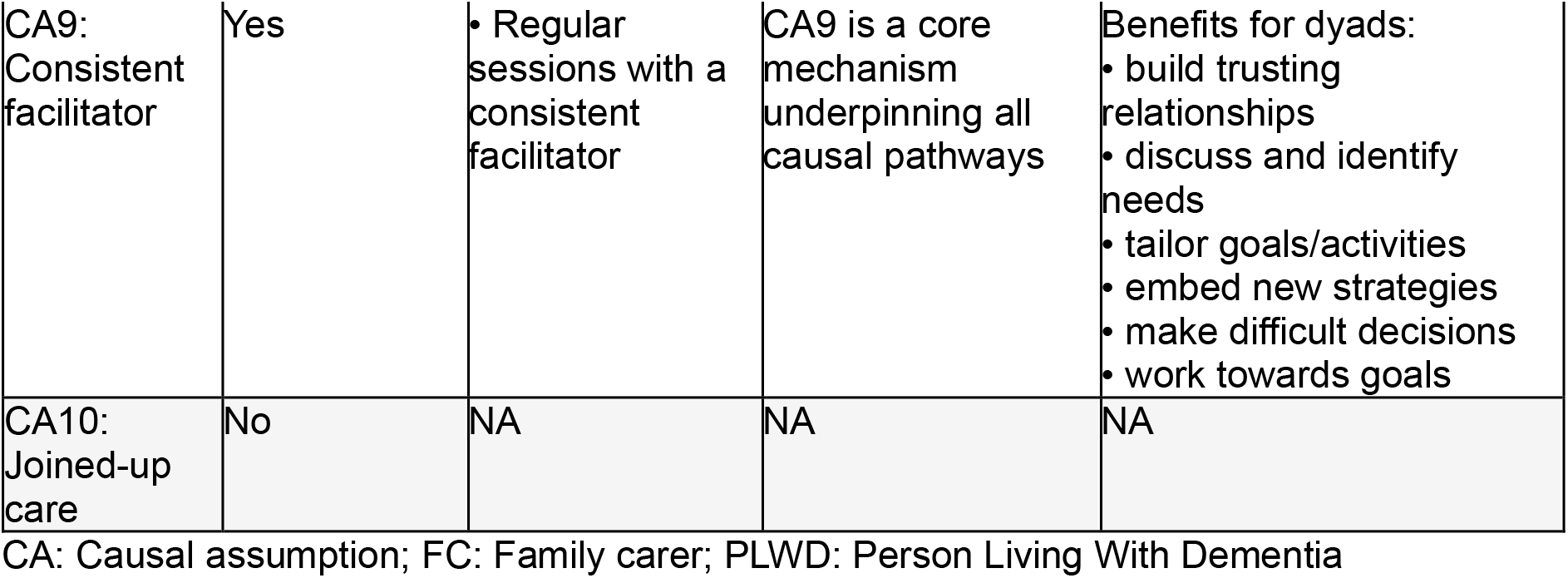
Integrated analysis summary.

The revised theory of change model (Figure 3) illustrates the updated causal assumptions, and how different causal pathways are important depending on dyadic engagement.

**Figure 3.**
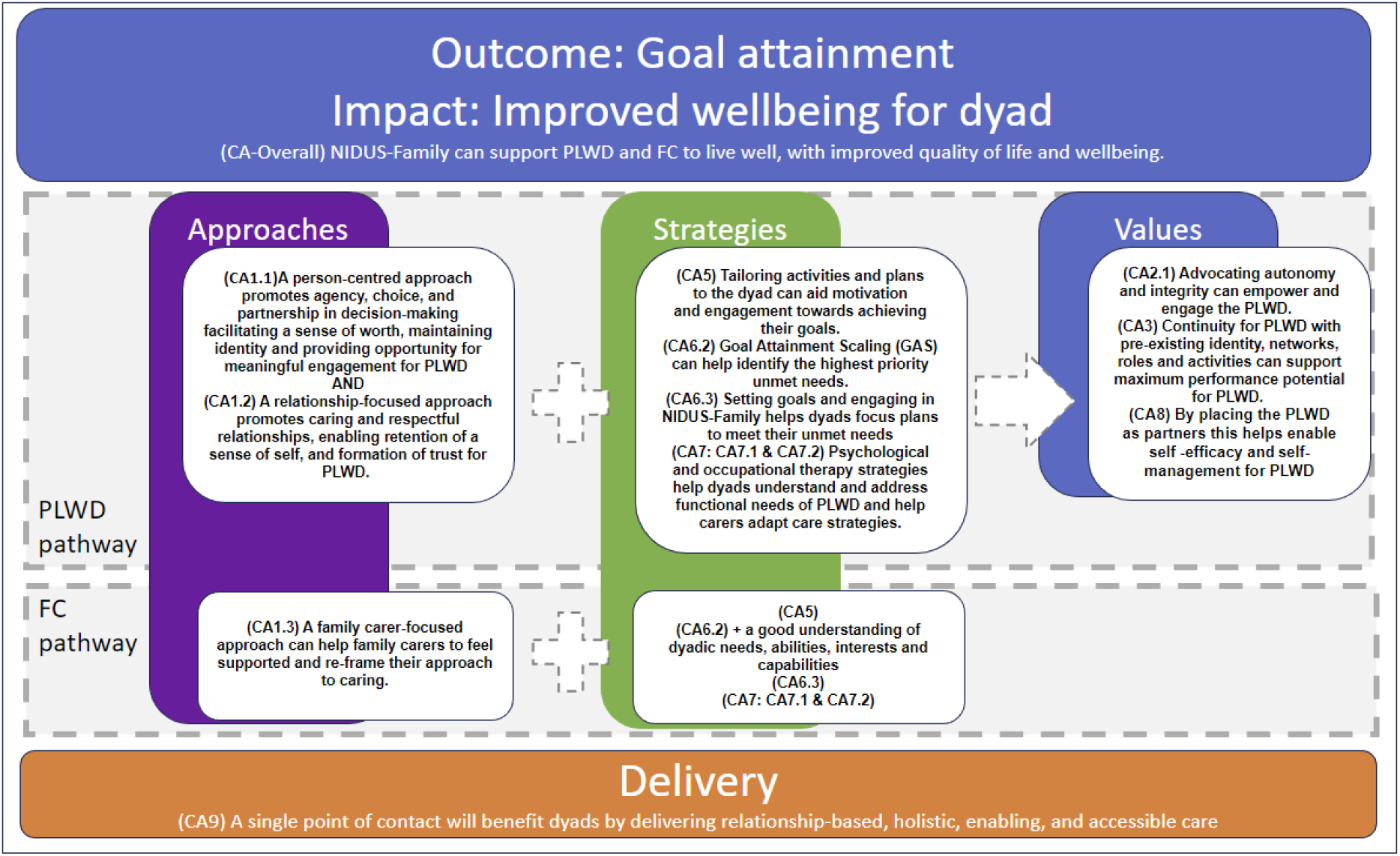
Revised theoretical model of change for attainment of dyadic goals through NIDUS-Family intervention

## DISCUSSION

Our findings show NIDUS-Family was delivered as intended. It worked through being relationship-, person- and carer-focused; supporting carers to take new perspectives, break down tasks into small steps and try out tailored approaches to personalising care with a consistent facilitator and dedicated space. Effects were realised through most pathways hypothesised in the theoretical model, with contexts determining which were most relevant. Where the care recipient was less engaged in care, pathways often focused on carer wellbeing and care strategies, and where they were more engaged, by enabling a person-centred approach.

The UK Dementia Guidelines recommend offering “*psychosocial and environmental interventions to reduce distress*”, personalised strategies for behavioural and sleep disturbance, and carer support (13). However, in dementia care: “*with few exceptions, proven interventions have not been translated for delivery in real-world settings*” (14). Positive outcomes in dementia care trials are rare. NIDUS-family, which is effective and designed to be scalable could transform care, if widely implemented.

Methodological strengths of this process evaluation are its theory-based design, and triangulated data sources (8,15). The recruited sample of people with dementia was balanced for gender, and the proportions from non-White ethnic groups, and who owned their home were broadly comparable to the older general population (16).

There were some limitations. Trials differ from real-world contexts (17). The intervention was only delivered in English, within a well-resourced team. Similar characteristics (being co-resident, married, the care recipient having capacity to consent) predicted completing the intervention and process evaluation participation; no dyads who withdrew from the intervention participated in the process evaluation, so the sample interviewed probably excluded those with less positive views about the intervention, indicated by their non-completion of it. The low acceptability questionnaire response rate may similarly have introduced bias. Though we interviewed people living with dementia where possible, our data primarily reflect a carer perspective.

This work highlights how Goal Attainment Scaling can be part of an intervention. Focusing on attainable goals is common in rehabilitation (18) and non-controversially helpful (19). Similar themes – allowing a shared language, and focusing on most meaningful outcomes – were noted in a goal-focused treatment programme for people with haemophilia (20). Understanding mechanisms behind how NIDUS-Family enable goal attainment will support its roll out into post-diagnostic care (21).

## Data Availability

All data produced in the present study are available upon reasonable request to the authors

## Funding

This work is part of the NIDUS (New Interventions in Dementia Study), which is hosted within the Alzheimer’s Society Centre of Excellence for Independence at Home (Centre of Excellence grant 330).

## Declaration of Interest

The authors declare no conflict of interest.

## Authors’ Contribution

All authors made a substantial contribution to this work. DW led the process evaluation, supervised by LTB, with SMT as an external advisor. DW and CC drafted the paper, which all authors revised for important intellectual content. JBu managed the study; VV and JBa were the trial statisticians. CC is the chief investigator of the study and secured its funding.

## Notes

### Competing Interest Statement

The authors have declared no competing interest.

### Clinical Trial

ISRCTN11425138; IRAS project ID: 271363

### Funding Statement

This work is carried out within the UCL Alzheimers Society Centre of Excellence (grant 300) for Independence at home, NIDUS programme

### Author Declarations

Ethical approval was received from Camden & Kings Cross Research Ethics Committee (REC). Study reference: 19/LO/1667

