## Supplementary figures and images for "Process evaluation of a New psychosocial goal-setting and manualised support intervention for Independence in Dementia (NIDUS-Family)"

### Supplementary Figure 1AS

**Figure 1AS: Observation checklist, responses to discrete items (n=12); values and approaches**

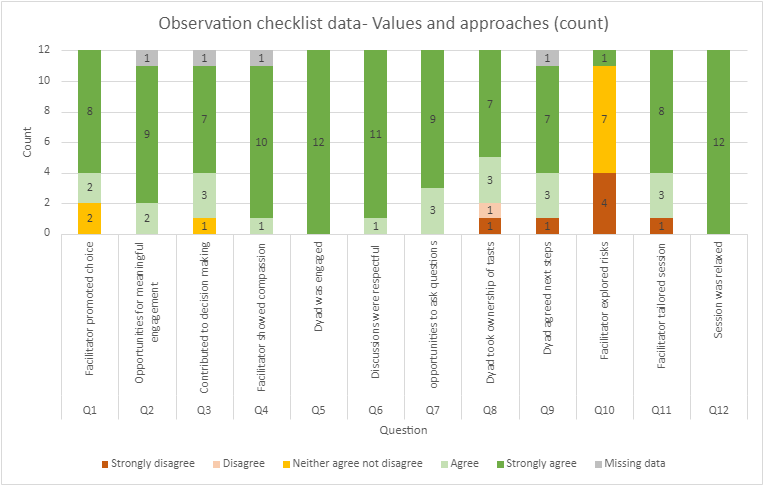
